# Formative Assessment and Cluster Membership of the Homeless Person’s ER Visits

**DOI:** 10.1101/2023.10.10.23296828

**Authors:** Gesulla Cavanaugh, Patrick Hardigan, Raymond Ownby, Stachyse Stanis, Prasanna Karur

## Abstract

The evidence is limited as to whether homeless individuals who visit Emergency Departments share similar characteristics with individuals from private households who, upon examination, are confirmed to need emergency health services beyond preventative healthcare. While the literature heavily addresses the homeless person in the ED, similarity and differences in characteristics with other social groups are lacking to guide the development of targeted health care and emergency health services for the homeless person. This explorative study investigates hospital ER data to draw inferences on ER patient characteristics and attempts to categorize patients according to residence. Nominal regression analyses reveal that cluster membership generated from ER data can predict patient residence and suggest that substance abuse and depression can predict 72-hour ER visit recurrence. Despite reporting low comorbidities, homeless patients were more likely to experience frequent ER visits within 72 hours and had higher rates of depression and substance abuse. Based on behavioral, health, and social characteristics of the homeless person, hospitals could consider targeted provisions and a follow-up mechanism to meet the health care needs of homeless patients.

## Introduction

The chronically homeless person has difficulty accessing the health care system appropriately and is inadequately equipped to navigate through available resources for disease management. Mismanagement of chronic disease, when combined with substance abuse, becomes exacerbated in the homeless person, and leads to detrimental outcomes. Continuous alcohol abuse is often one of the reported factors that retains individuals in homelessness and is associated with unintentional injuries, life threatening intoxication, and is one of the key factors linked to the homeless person’s abuse of the Emergency Department [1,2]. Evidence from the literature asserts that the homeless population has a higher rate of alcohol and substance abuse than housed adults [3,4,5] and because of the homeless person’s constant exposure to harsh social and untiring conditions, their inability to live a healthy lifestyle is even further multifaceted.

Although U.S. population estimates point to a continuous decrease in homelessness nationally, in 2018 there were 552,830 homeless individuals, which was a 0.30% increase from 2017 [6]. Of this number, 358,363 were sheltered, and 194,467 were unsheltered [6]. The U.S. experienced a small increase in homelessness from 2017 to 2018 where unsheltered homelessness increased by 2% [6]. The U.S. Department of Housing and Urban Development (2018) reported that there was a decrease in overall homelessness for all racial groups except for Caucasian which was shown to experience a 4% increase in homelessness [7]. Unsheltered homelessness increased among people who identified as White (8%), Asian (2%), and Multiracial (8%) [7]. Meanwhile, African Americans were highly represented in the sheltered group, marking up 47% of the sheltered homeless, but only 26% of the unsheltered group [7]. From 2019 to 2020, the homeless population increased by 2.22% [6]. However, from 2020-2021, during the first year of the COVID-19 pandemic, a reported 41.59% decrease in homelessness was recorded [6]. Different factors are attributed to this significant decrease as many reporting resources such as Point-in-Time counts were disrupted due to the pandemic. Research within the context of the growth in homelessness, its associated health challenges, and its wide health disparity suggests that homeless individuals are one of the ineffectively treated groups in the Emergency Room departments mainly due to the behavioral health factors mentioned above [5].

Individuals from the homeless population are one of the leading groups misusing Emergency Room (ER) services in many highly populated cities in the U.S. [3,8]. ER visits from individuals experiencing homelessness are usually recurrent in which these individuals may present with a combination of and worsened health and social conditions (i.e., depressed, intoxicated, and hypertensive) [3]. Those over the age of 50 who are homeless and who visit the ER are more likely to arrive in critical conditions, and subsequently, have a higher risk of mortality [7,8]. Additionally, the needs of the homeless person are not clearly recognized in the Emergency Room Department processes [8]. While homeless patients utilize the ER at a different rate than housed adults, their complex needs for attending the ER may not be met by the procedure of existing services and processes in the hospital [8, 9]. Consequently, they are often discharged without receiving targeted health education and the appropriate resources to manage their conditions [8, 9].

To examine the characteristics of individuals who visited the ER of selected hospitals in the United States at a given time point, this study employed an explorative and machine learning analytical approach with particular focus on the homeless person. The aims of the analyses were to assess if any significant differences exist between individuals who visited the ER based on residence type (private residence, nursing home, homeless). Furthermore, because recurrent and preventative ER visits are consistently problematic and heavily affect hospital operation, we aimed to determine if there were significant differences in patient characteristics who had more than one ER visit within 72 hours, and whether factors such as residence type, perception of pain, depression, and ER visit urgency could predict recurrent ER visits within 72 hours. The hypotheses for this explorative study are as follow:

H1o: There is a significant relationship between recurrence of ER visits and residence type, perception of pain, depression, and ER visit urgency.

H2o: Significant differences in ER patients’ characteristics exist based on cluster formation.

## Materials and Methods

### Hospital Database

Published Hospital data were leveraged from the Ambulatory Health Care Data, made possible by The National Health Care Surveys which is comprised of the National Hospital Ambulatory Medical Care Survey (NHAMCS) [10]. The NHAMCS has been leading in assessing ambulatory medical care in hospital emergency departments since 1992. The database is composed of yearly sample records from approximately 500 nationally representative hospitals within the United States where the Emergency Department sample data are collected during a 4-week reporting window and may vary depending on the hospital. NHAMCS data help yield meaningful descriptive statistics pertaining to the demographics, treatment, medical conditions, diagnostic services rendered, and medication prescribed for those who visit the hospital emergency departments (ED). The NHAMCS was selected for its robustness on sampling patients’ activities within the ED. For analysis pertaining to this explorative examination, Emergency Room visit data were analyzed for 2013-2016 from the NHAMCS database. This comprised records on 89,149 patients; patients with unknown residence were omitted from the analyses which yielded a total of 85,830 persons, classified for the purpose of this study as having either a private residence, living in a nursing home, or homeless (N= 83,446; N= 1459; and N= 925, consecutively).

### Statistical Analysis

To address whether residence type could predict 72-hour ER visit recurrence and the characteristics of the individuals visiting the ER, a series of analyses were performed using JMP v16.0 and SPSS Modeler v18.1. Data from 2013-2016 were investigated and in-depth analyses were completed on the 2016 database for its inclusion of more study-specific variables. The 2016 database, composed of records on 19,467 patients, housed more relevant data and comprised more pertinent variables relating to the aims of this study; as such it was selected to address the study hypotheses. 650 patients from the 19,467 had unknown residence and were excluded from the analyses.

Additional statistical analyses were conducted to investigate the relationship between residence type, perception of pain, depression, and ER visit urgency, and assessed if the selected variables predict 72-hour ER visit recurrence. Because the Generalized Regression procedure fits GLM with a process of penalization as a process to control for overfitting, it was selected for its optimized model estimation capability to assess the most appropriate model for 72-hour ER visit recurrence; a stepwise regression analysis was conducted following the GLM method.

To address the second hypothesis and identify patient grouping characteristics, four-step cluster analyses were applied with different variables. For the analyses involving cluster membership, a two-phase process was completed. In the first phase, we used k-means clustering algorithm to generate three clusters based on the Elbow method [11] for ER patients constructed from the selected variables, total co-morbidities, 72-hour ER visit recurrence, and substance abuse or dependence. The generated cluster result was saved as a variable assigning a cluster membership for each patient in the database. In the second phase, a nominal logistic regression was applied with cluster membership used as an outcome variable with residence type as a possible predictor.

## Results

### Behavioral, Health, and Social Characteristics

For 2013-2016, 25% (n= 364) of those living in a nursing home reported having Alzheimer’s Disease or Dementia in comparison to 0.4% (n= 4) in the homeless group and 0.9% (n= 719) in the private household group. Nonetheless, 30% (n= 277) of the those classified as homeless reported having alcohol misuse, abuse, or dependence, in comparison to 2.9% (n= 2,419) and 1.4% (n= 20) from those from private homes and those in nursing homes. Twenty-six percent of those classified as homeless reported having depression, in comparison to 21% from those in nursing homes, and 9% from those in private households.

In the 2016 database composed of 272 homeless patients, 287 nursing home patients, and 18,258 patients from private residence, further results indicate that patients from the homeless group (*M*= 0.126, *SD*= 0.33) were more likely to have recurrent ER visits within 72 hours (no, yes), compared to those from private homes (*M*= 0.031, *SD*= 0.172), and those from nursing homes (*M*= 0.019, *SD*= 0.139) (see table 1 and figure 1). Sixty nine percent of the homeless group were males, but this was the opposite in the private residence group (45%) and the nursing home group (45.3%). Diagnoses of Alzheimer’s Disease/Dementia and cardiovascular disease were higher in the nursing home cohort, which is expected as the mean age for that group was higher. The occurrence of congestive heart failure was 2.7% for the private residence group, 18.5% for the nursing home group, and 1.8% for the homeless group; cerebrovascular disease/ history of stroke or transient ischemic attack was 2.8%, 19.2%, and 2.6%, consecutively; and Alzheimer’s Disease/Dementia was 0.9%, 26.5%, and 0.7%. The mean age for the private residence group was 37.27 (*SD*= 23.70), the nursing home group was 73.34 (*SD*= 18.74), and the homeless group was 43.10 (*SD*= 14.61). The distribution of recorded blood pressure was different between the homeless (*M*= 81.45, *SD*= 15.12), and those from private residence (*M*= 78.69, *SD*= 14.74) and the nursing home group (*M*= 73.36, *SD*= 15.71), highlighting higher values for the homeless population (figure 2), with the results persisting when stratified by age (figure 3).

**Table 1.** Descriptive statistics by residence type.

|  | Private Residence (n= 18,258) |  | Nursing Home (n= 287) |  | Homeless (n= 272) |  |
| --- | --- | --- | --- | --- | --- | --- |
|  | M (SD) | % | M (SD) | % | M (SD) | % |
| 72-hr ER | 0.031(0.172) | — | 0.019(0.139) | — | 0.126(0.33) | — |
| AD | — | 0.9 | — | 26.5 | — | 0.7 |
| BP-Dias | 78.668(14.737) | — | 73.359(15.71) | — | 81.453(15.127) | — |

**Figure 1.**
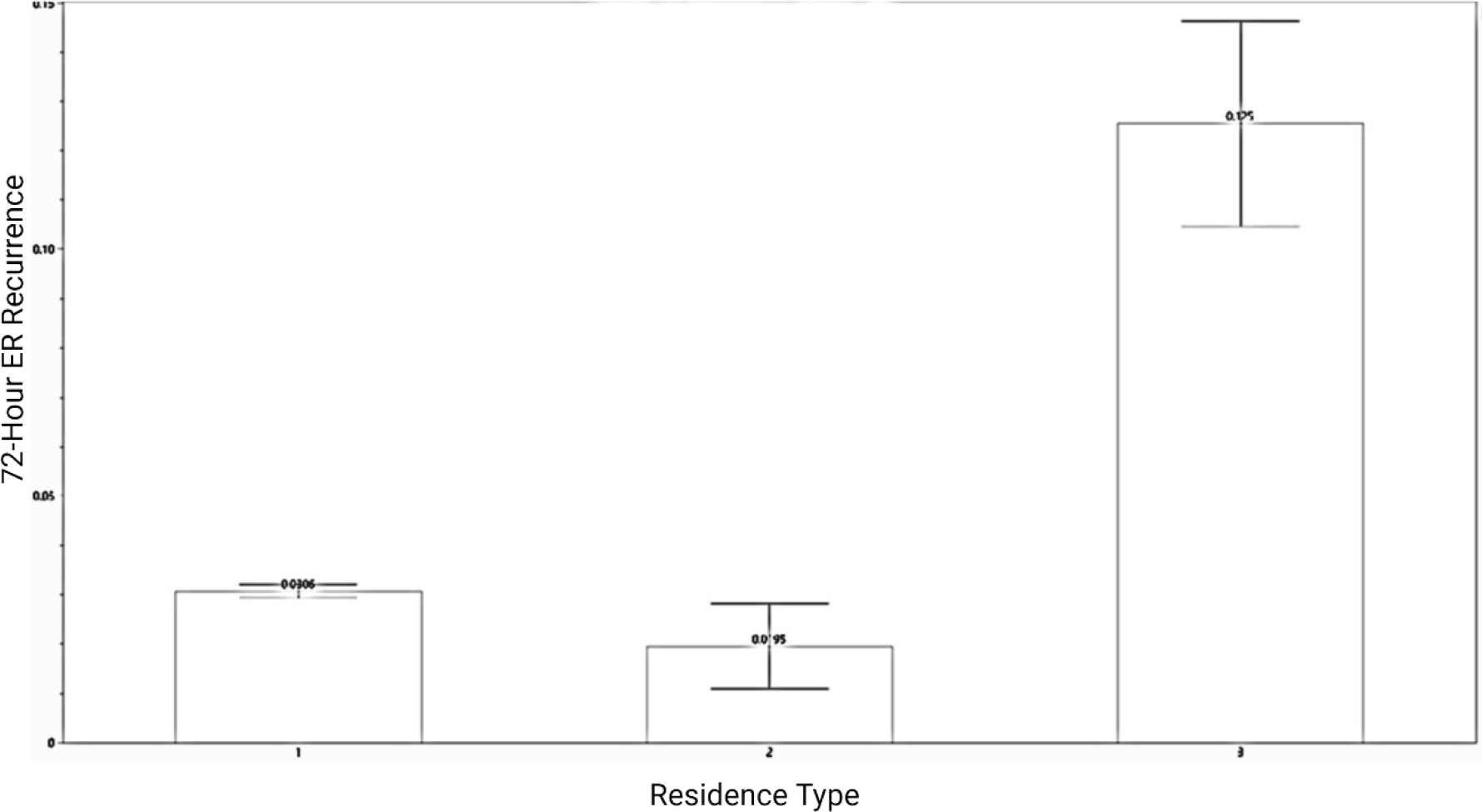
Mean ER Visits within 72 Hours by Residence Type

**Figure 2.**
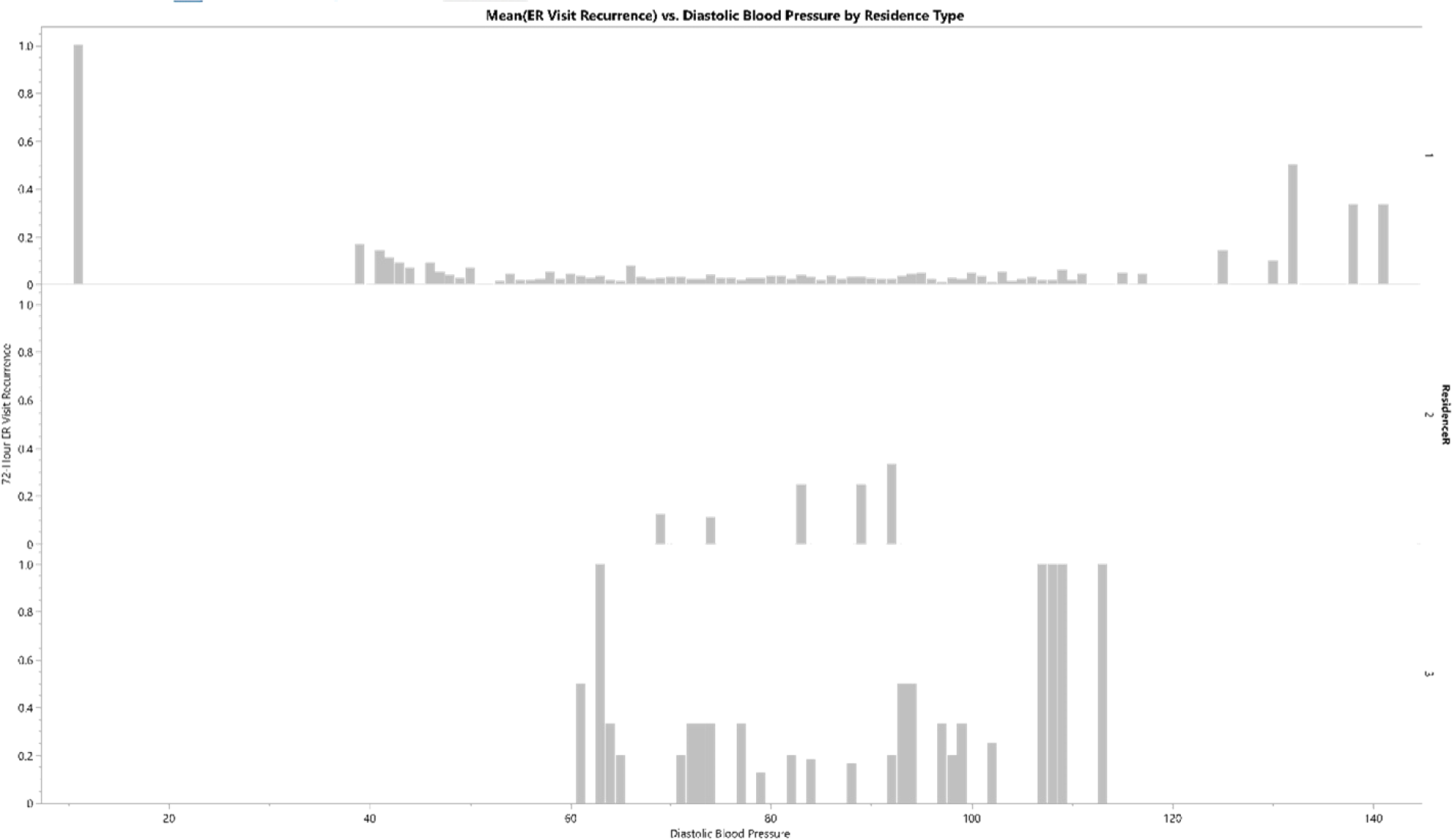
Mean ER Visits within 72 Hours vs. Diastolic Blood Pressure by Residence Type

**Figure 3.**
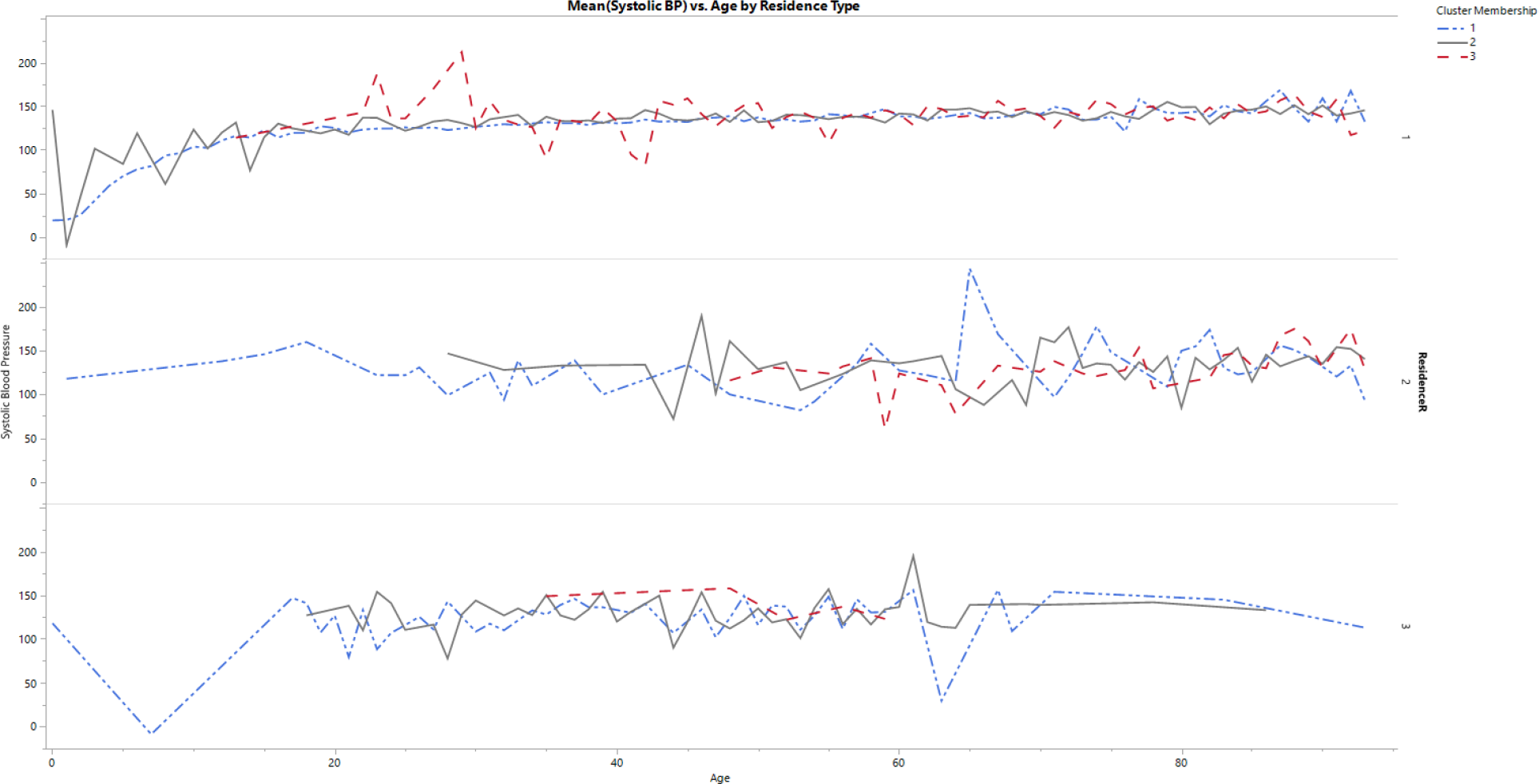
Mean Systolic Blood Pressure vs. Age by Residence Type

For the given 2016 dataset, five percent (n= 921) of the patients from private residence reported substance abuse or dependence, compared to 1.5% (n= 5) from the nursing home group and 34.2% (n= 93) from the homeless group. The proportion of patients who reported substance abuse or dependence differed significantly by residence type, *X*^*2*^ (2, n= 18,817) = 452.19, *p* < .001. Analyses of variance (ANOVA) on residence type yielded significant variation with substance abuse, *F*(2, 18,814) = 214.716, *p* < .001; and with depression, *F*(2, 18,814)= 62.648. Post hoc Tukey HSD tests showed that for substance abuse, the private residence group did not significantly differ from the nursing home group (*p*= 0.961) but was significantly different from the homeless group (*p* < 0.001); the nursing home group was significantly different from the homeless group (*p*< 0.001). For depression, the private residence group differed significantly from the nursing home group (*p*< 0.001) and was significantly different from the homeless group (*p*< 0.001); the nursing home group was not significantly different from the homeless group (*p*= 0.535).

The results from the stepwise regression analysis suggest that with 72-hour ER visit recurrence as the outcome variable, residence type, perception of pain, depression, and ER visit urgency predict a significant amount of criterion variance. The model was significant *F*(4, 17,818) = 5.498, *p<* 0.0002, R^2^= .0012. Residence type was the most significant contributor to the model (t= 3.84, *p*< 0.0001), depression was the second significant contributor (t= 2.10, *p<* 0.036), while perception of pain and urgency to be seen did not contribute significantly to the model (t= 0.56, *p*= 0.578; t= 1.21, *p*= 0.221).

### Cluster Formation and Membership Profiles

Table 2 shows results of the three-cluster solution generated from three variables selected for their implication in the study aims. Each cluster is composed of a combination of classifications from the input variables. This clustering process designates the variable with the highest mean in a respective cluster as the identify of that cluster (table 2). In this regard, for example, in cluster 3, comorbidity has a high mean score of 5.844, and as such, that cluster is heavily represented by the co-morbidity variable. In the cluster solution, the first cluster (cluster one) is mainly composed of patients with no substance abuse or dependency, with low comorbidities, and those who did not have recurrent ER visits in the past 72 hours. The second cluster contains patients who had some comorbidities, with substance abuse and dependency, and with diverse members who did or did not have recurrent ER visits within the past 72 hours. The third cluster is dominated by patients with multiple comorbidities, and some members with substance abuse or dependency and who were more likely to have recurrent ER visits within the past 72 hours than the other two clusters (figure 4).

**Table 2.** Clusters with Co-Morbidity, Substance Abuse, and ER Visit Recurrence.

|  | **1** (n=12,727)<br>**M(SD)** | **2** (n= 4,192)<br>**M(SD)** | **3** (n=782)<br>**M(SD)** |
| --- | --- | --- | --- |
| 72-Hour ER Visit | 0.035(0.185) | 0.017(0.129) | 0.056(0.230) |
| Substance Abuse | 0.000(0) | 0.152(0.359) | 0.015(0.123) |
| Co-Morbidity | 0.288(0.462) | 2.618(0.857) | 5.844(1.287) |
| Residence Type | Count | Count | Count |
| 1_ | 12208 | 3762 | 690 |
| 2_ | 65 | 135 | 55 |
| 3_ | 124 | 118 | 7 |

**Figure 4.**
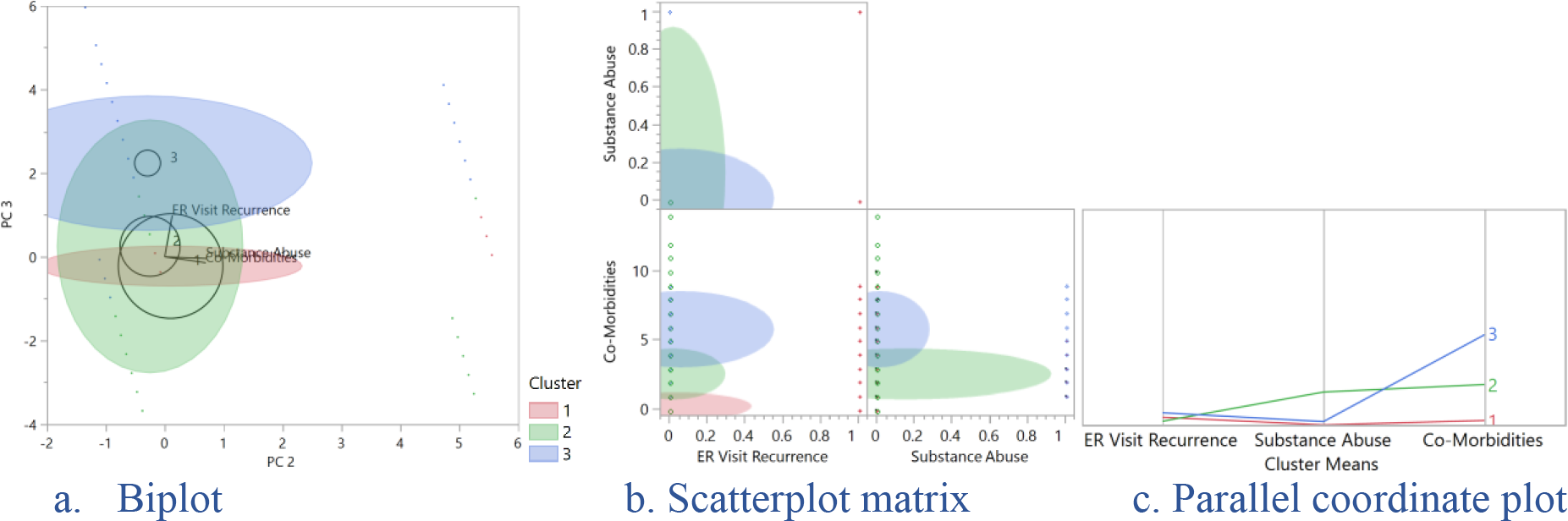
Cluster Membership Comparison

To identify if residence type was associated with cluster membership, a nominal logistic regression was performed using cluster membership as the outcome variable. Analyzing the association between residence type and cluster membership revealed a statistically significant relationship *𝒳*^*2*^(4, *n*= 17,164) = 338.16, *p* < .0001. The model explained 1.4% (Rsquare (U)) of the variance in cluster membership, indicating higher uncertainty accounted for the model; adding other variables of interest such as depression and diastolic blood pressure to the model did not significantly improve the proportion of uncertainty (data not shown). Nonetheless, regardless of uncertainty, the cluster analyses including only some variables of interest, provided a frame of assembly on ER patient characteristics based on residence type.

Following the analyses, the Prediction Profiler was used in JMP to assess likely predictors and determine the robustness of the generated model. The model suggests that homeless patients had a 0.47 probability to belong in cluster three, indicating a higher likelihood for membership in that cluster than the other two clusters (0.23, 0.30); patients from nursing homes had a 0.65 probability to belong in cluster 3. Members of cluster one were more likely to be patients from private residence (0.73), while cluster two has equal likelihood to contain patients from all three residence types (Figure 5). In comparing cluster membership with associated covariates, including total chronic illnesses, pulse rate, and age, the results further reveal that cluster three is composed of mainly older individuals with more comorbidities, with higher membership counts in clusters two and three (figure 6).

**Figure 5.**
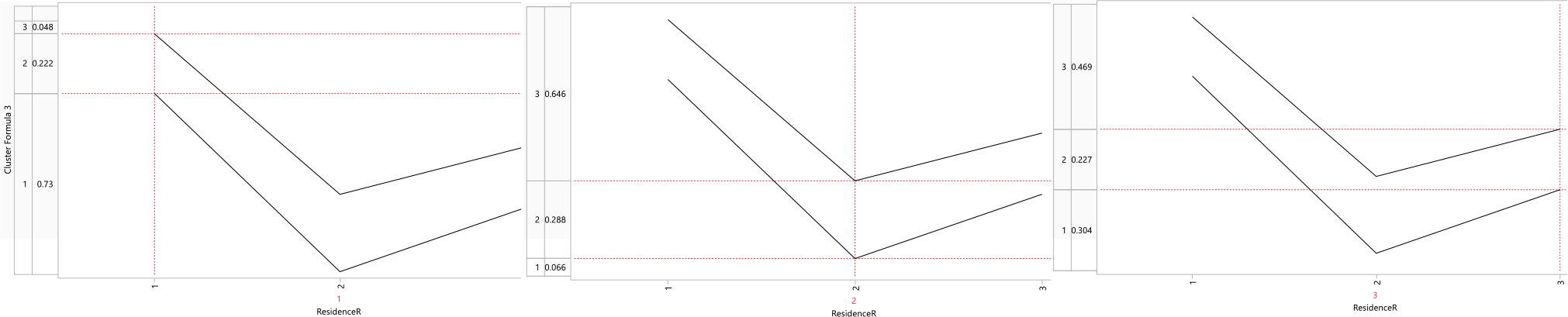
Prediction Profiler for Cluster Membership vs Residence Type

**Figure 6.**
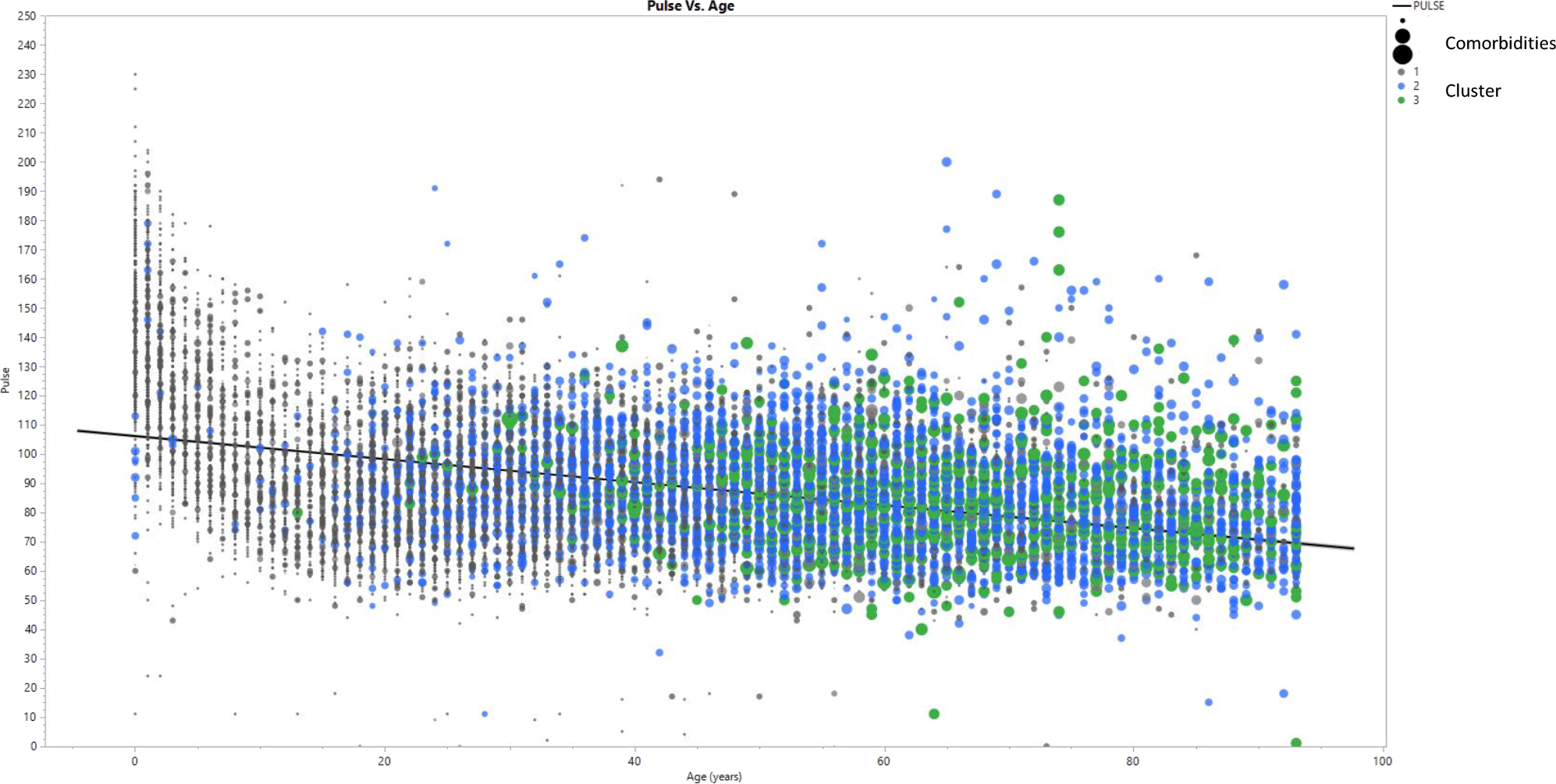
Cluster membership and Relationship with Covariates

## Conclusion

The results from the analyses showed that the homeless person who visits the ER has significantly higher levels of substance abuse and dependency and more depression compared to those living in private homes and nursing homes. Moreover, challenging conditions may force individuals from different residences to visit the ER on multiple occasions. The results suggest that it is possible to categorize ER patients based on several factors, including residence type, co-morbidities, and depression; nonetheless, there is a challenge to truly separate the homeless patient from the other groups based on only health variables, hence why knowledge of the patients’ residence type is crucial. The results are consistent with previous findings, such as Hopkins and Narasimhan (2022) who claim that homelessness, coupled with substance abuse and mental illness, aggravates disease burden and early biophysiological deterioration in individuals, and increases the probability of using the emergency department.

We had hoped to find stricter margins for subsets of individuals with different health and social conditions based on residence type. Nonetheless, using cluster analysis, we were not able to generate a distinct cluster membership for the homeless patients who visit the ER. This indicates that the homeless persons who present to the ED may have similar health and social characteristics as the other groups, with no significance within group variance. Moreover, the results in general suggest that although the homeless person and the general population have similar health characteristics, the homeless person has a 72-hour ER visit recurrence at higher rates than individuals from private residence. The homeless person may, instead, visit the ED when health issues become unbearable, and as a last resort. Future analyses will take within group characteristics into context as applicable to only the homeless person, examining innovative solutions for addressing this group and its presented health challenges.

Overall, the results of these analyses suggest that cluster analyses have potential to reveal social health and group characteristics and can support targeted solutions respective to group individualities in the ER. The formative assessment of the homeless individual suggests that they may be utilizing the ED more frequently than individuals from the general population. To mitigate this problem, potential solutions include focusing on providing educational information to the homeless patient and encouraging connections to a social worker who may mitigate the circumstances, thereby alleviating worsened health conditions and reducing preventative ER visits.

## Data Availability

All data produced in the present study are available upon reasonable request to the authors

https://www.cdc.gov/nchs/ahcd/nhamcs_participant.htm

